## supplementary for "Testing and isolation to prevent overloaded health care facilities and to reduce death rates in the SARS-CoV-2 pandemic in Italy"

RESEARCH

### Additional file 1

Arnab Bandyopadhyay*[1†], Marta Schips*[1†], Tanmay Mitra[1], Sahamoddin Khailaie[1], Sebastian C. Binder[1] and Michael Meyer-Hermann[1,2,3*]

*Correspondence:



[1]Department of Systems Immunology and Braunschweig Integrated Centre of Systems Biology (BRICS), Helmholtz Centre for Infection Research, Rebenring 56, 38106, Braunschweig, Germany
Full list of author information is available at the end of the article
†These authors contributed equally to this work (alphabetical order).^These authors contributed equally to this work (alphabetical order).

**Estimation of undetected cases:** Supplementary text with detailed description on IFR estimation method.

**SECIRD models:** Supplementary equations of the models used in the analysis.

**Identifiability analysis:** Supplementary text with detailed description on parameters' identifiability analysis and its results.

**Figure S1:** Excess Deaths.

**Figure S2:** Age-specific IFR.

**Figure S3:** MCMC results.

**Figure S4:** Impact of the testing frequency on the regional epidemics.

**Figure S5:** Structural identifiability test.

**Figure S6:** Practical identifiability.

**Figure S7:** Fit up to July 23$^{rd}$.

**Figure S8:** $\mathcal{R}_t$ curves.

**Figure S9:** $\mathcal{R}_t$ sensitivity.

**Figure S10:** Impact of testing upon $\mathcal{R}_t$ evolution.

**Figure S11:** *Capacity* model fit.

**Figure S12:** Impact of 5 times more tests.

**Figure S13:** Variation of excess dead results depending on $\alpha$ values for Lombardia.

**Keywords:** SARS-CoV-2; COVID-19; Epidemiology; Modeling; Non-pharmaceutical interventions; Reproduction number; Healthcare usage; Italy; Lombardia; Veneto;

### Estimation of undetected cases

*Data preprocessing*

Demographic and mortality data are available from the Italian Institute of Statistics' (ISTAT) website [1, 2]. ISTAT collects mortality data from the Italian *National register office for the resident population* (ANPR). Daily deaths from 2015 to 2019 are available stratified by gender and age. For 2020, data are available up to April 15$^{\text{th}}$, listing 6866 municipalities and covering 86% of the Italian population. For each region, the municipalities list include those for which the death data set is complete and we, accordingly, extracted demographic data for these municipalities. Here, we considered seven age groups: 0-20, 21-40, 41-50, 51-60, 61-70, 71-80 and 81+. For each region and for each age range, we counted the daily deaths for the period January to April 15$^{\text{th}}$ 2015-2020. For each region, we then summed up daily deaths of municipalities by age groups. Similarly, from the demographic data we calculated the population size of each age group. Due to the unavailability of demographic data for 2020, we used 2019 data as a proxy of 2020. To calculate the Infection Fatality Rate (IFR) for Italy, we considered mortality and demographic data of municipalities across all regions for which data are complete and followed the same procedure.

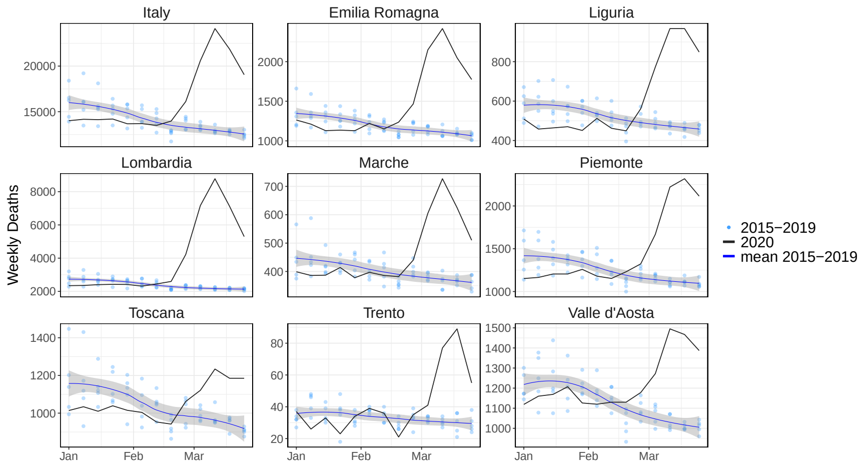


**Figure S1 Excess Deaths.** Weekly deaths for the period of January 1$^{\text{st}}$ to April 15$^{\text{th}}$ for 2015-2020 of different regions in Italy. Data are acquired from ISTAT and include 6866 municipalities, covering 86% of Italian population. Light blue points: all-cause deaths in 2015-2019; blue line with grey shaded area: spline fit of 2015-2019 deaths; black line: all-cause deaths in 2020.

*Bayesian estimation of COVID-19 IFR*

To estimate IFR, we implemented maximum likelihood estimation for the binomial distribution of the mortality rate under a Bayesian framework. Our model is adopted from [3] and modified accordingly. The model assumes that the observed number of deaths in the COVID-19 period (February 20$^{\text{th}}$ to April 15$^{\text{th}}$) of each year is binomially distributed according to:

$$
\begin{aligned}
D_{\mathrm{a,y}} &\sim \mathrm{Binomial}(\delta_{\mathrm{a}}, N_{\mathrm{a,y}}) && \text{for y } \in [2015, 2019] \\
D_{\mathrm{a,y}} &\sim \mathrm{Binomial}(\delta_{\mathrm{a}} + \delta_{\mathrm{a}}^{\mathrm{Covid}} \cdot \theta, N_{\mathrm{a,2019}}) && \text{for y } = 2020
\end{aligned}
$$

where $a$ denotes the seven age groups, mentioned in the previous section and $y$ denotes the year. $D_{\mathrm{a,y}}$ and $N_{\mathrm{a,y}}$ denote total death and population of age $a$ in year $y$, respectively. $\delta_{\mathrm{a}}$ is the baseline death rate of age $a$ and is heterogeneous across age groups. To model deaths in 2020, in addition to $\delta_{\mathrm{a}}$, we considered a COVID-19 death rate, $\delta_{\mathrm{a}}^{\mathrm{Covid}}$ multiplied with the exposed fraction, $\theta$. $\delta_{\mathrm{a}}^{\mathrm{Covid}}$ is the IFR of age $a$ and it is assumed to be absent in the previous years. $\delta_{\mathrm{a}}^{\mathrm{Covid}}$ is sampled from a uniform distribution with range between 0 and 0.2. IFR is less than the Case fatality rate (CFR) if and only if the detected cases are more likely to lead to death than undetected cases, which is indeed the case for COVID-19. For COVID-19, the reported CFR is within the range of 5% to 15% and several estimation methods resulted IFR well below 20% including the 70+ age group [4, 5, 6, 7, 8, 9]. Therefore the upper bound for IFR has been chosen to 0.2. $\theta$ is the infection rate (IR) or attack rate and denotes the fraction of the population that is exposed. As supported by a recent seroprevalence study [10] it is assumed to be constant across all age groups. Moreover, seroprevalence studies indicate that population-wide immunity is, in general, less than 50% [11, 10, 12], thus we sampled $\theta$ from a beta distribution for which the density peaks between 20% and 40%. In a nutshell, we are estimating fifteen parameters (considering seven age groups), $\delta_{\mathrm{a}}$, $\delta_{\mathrm{a}}^{\mathrm{Covid}}$, and $\theta$ from the observation of death data of previous years (2015-2020) classified by age groups (42 data points) given the age distribution of population (2015-2020, 42 values).

We used the following priors to estimate the $\delta_{\mathrm{a}}$, $\delta_{\mathrm{a}}^{\mathrm{Covid}}$ and $\theta$:

$$
\begin{aligned}
\delta_{\mathrm{a}} &\sim \mathrm{Uniform}(0, 0.1) \\
\delta_{\mathrm{a}}^{\mathrm{Covid}} &\sim \mathrm{Uniform}(0, 0.2) \\
\theta &\sim \mathrm{Beta}(3, 5)
\end{aligned}
$$

For each region, the model was evaluated using the Markov Chain Monte Carlo (MCMC) sampling method. We used 30 independent chains; each drew 50,000 samples from the joint posterior distribution. We discarded the first 5000 as burn-in periods from each chain. We merged the results of all 30 independent chains and calculated the 95% credible interval by using the 95$^{\mathrm{th}}$ quantile of the posterior distribution. Posterior distribution of parameters are shown in Fig. S3. The total number of infections for each region was calculated from the definition of IFR, which is death over total infection. Here we considered the number of deaths as the excess deaths of February 20$^{\mathrm{th}}$ to April 15$^{\mathrm{th}}$ with respect to the average of the previous years (2015–2019). Simulation results are reproducible and were carried out using R version 3.6.2 [13, 14, 15, 16].

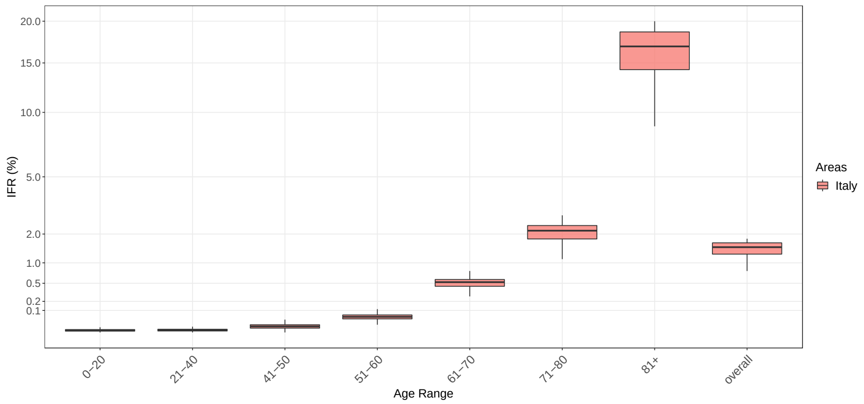


**Figure S2 Age-specific IFR.** Boxplot of estimated IFR across the age ranges and of overall IFR, for Italy and different regions. IFR is estimated by Bayesian MCMC framework (see Methods). IFR is significantly higher in the 81+ age group.

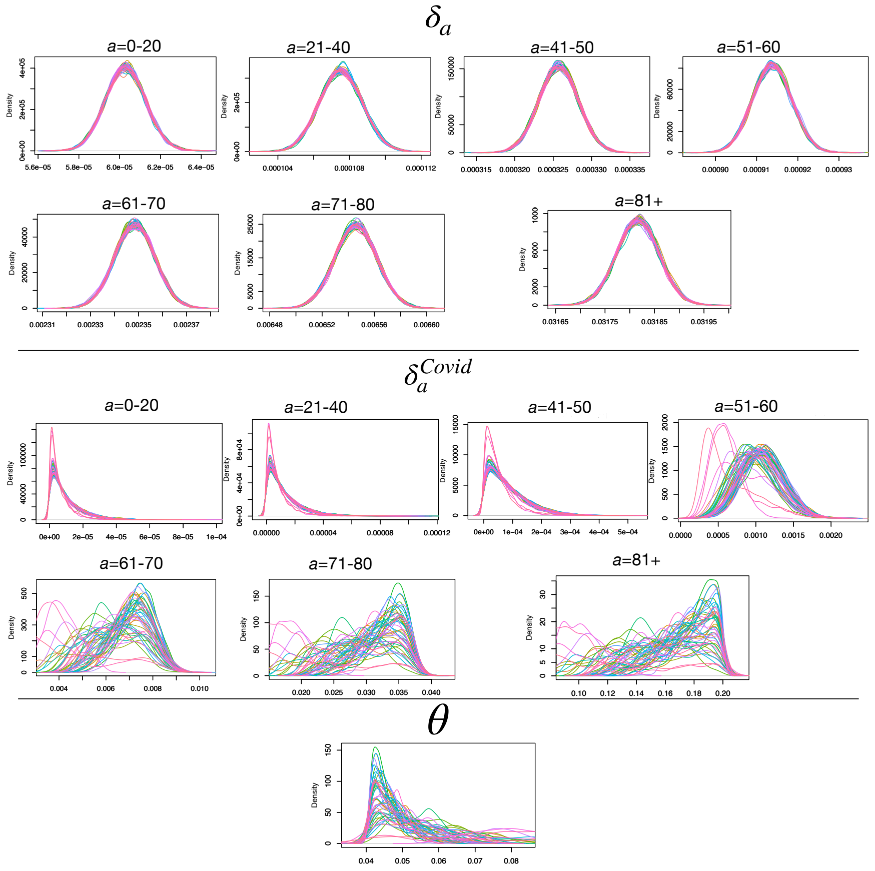


**Figure S3 MCMC results.** Posterior distribution of baseline lethality rates ($\delta_{\mathrm{a}}$), IFR ($\delta_{\mathrm{a}}^{\mathrm{Covid}}$) for seven age groups, and Infection Rate ($\theta$) of 30 independent MCMC chains. To estimate $\delta_{\mathrm{a}}$, $\delta_{\mathrm{a}}^{\mathrm{Covid}}$, $\theta$, we used following priors: Uniform(0,0.1), Uniform(0,0.2), Beta(3,5).

*Estimation of regionwise infection fatality rate (IFR)*

The severity of an epidemic can be characterized by the case fatality rate (CFR), defined as the percentage of deaths among the total number of diagnosed infections, and has been of high interest since the very beginning of the COVID-19 outbreak [17, 18]. On the other hand, infection fatality rate (IFR) is defined as the percentage of deaths among all infections, including the undiagnosed infections. For COVID-19, the true number of cases is unknown as a substantial portion of the infections are either asymptomatic or mildly symptomatic and remained undetected [19, 20]. At the beginning of the pandemic, testing was limited only to the symptomatic patients due to clinical findings suggesting that symptomatic cases are the major source of disease spreading [21, 22, 23, 24, 25]. It is, therefore, very difficult to get a reliable estimate of the true number of infections. A few methods have already been proposed to quantify the true number of infections and, hence, a realistic IFR [4, 26, 5].

To estimate undetected infections amid the COVID-19 pandemic, we analyzed the mortality rate of previous years and deaths in the year 2020. Demographic and death data of the Italian regions have been collected from the Italian Institute of Statistics (ISTAT). According to the report published by ISTAT, from the first COVID-19 death in Italy (February 20$^{th}$ 2020) to March 31$^{st}$ 2020, excess death in 2020 was 25,354 (total 90,946) compared to the previous five years (65,592 as average of 2015-2019). 54% of the additional deaths were diagnosed as COVID-19 positive [27]. In Fig. S1 we show region-wise, weekly deaths from January 1$^{st}$ to April 15$^{th}$ 2020. For the Italian regions where the pandemic started, like Lombardia, Veneto, Piemonte, the observed mortality of this year was substantially higher than previous years. For these regions, we estimated the total infection and associated IFR by implementing a Bayesian framework by adapting a standard binomial model (see previous section and [3]).

After the first identification of a COVID-19 case on February 20$^{th}$ 2020 in Codogno Hospital, near Lodi, Lombardia [28], the number of reported positive cases increased to 36 in the next 24 hours and, interestingly, the new cases were not linked to the first case, suggesting that the virus was circulating before its first identification. This is reflected in our estimation of undetected cases (Table 2). High increase in death over this year was observed in some cities, like Bergamo (568%), Cremona (391%), Lodi (371%), Brescia (291%), Piacenza (264%), Parma (208%) [27]. In the northern regions of Italy, especially where the initial outbreak occurred, for example in Emilia Romagna, Piemonte and Veneto, undetected infections were nearly 10 fold higher than the reported cases; and in Lombardia more than 21 fold. We observed substantial heterogeneity of the IFR across different age groups. For the age ranges below 60, it was determined as low as 0.05%. IFR was substantially higher in the 81+ age group (9.5% to 20%, Fig. S2). Despite Italy having the highest COVID-19 deaths in Europe, estimated infection rates (IR) were relatively low (highest in Lombardia $\sim$ 13%) across all regions, and hence the population was far from reaching the herd immunity threshold ($\sim$ 70%, assuming no previous immune memory). Our estimated total infections (detected + undetected) are close to the numbers reported in [29].

*Correlation between undetected cases and test frequency*

Despite being hit hardest by COVID-19, some of the Italian regions handled this crisis situation better and managed to contain the virus. For instance, in Veneto, CFR was 6.4%, 3 fold lower when compared to Lombardia at 18.3%. This is also reflected in the IR, 2.61% in Veneto while in Lombardia it was 13% despite their geographical proximity. The testing strategy implemented by these two regions was completely different. Most regions, like Lombardia and Piemonte, followed the World Health Organization (WHO) and central health authority indications by mainly testing the symptomatic cases, while Veneto implemented a much more extensive population testing. Toscana followed a testing strategy very similar to Veneto by ramping up its testing capacity quickly. To determine whether implementing different testing strategies succeeded in keeping the undetected and the overall infection amount under control, we investigated the association between the rate of tests performed by regions and total infections (Table 2) in the early phase of the pandemic. We measured the Kendall and Distance correlation between the total infections (up to April 15$^{\text{th}}$ and including undetected infections) normalized by the population size and the total tests performed (up to April 15$^{\text{th}}$) per reported infection. This yielded a significant correlation with a coefficient of -0.67 and 0.81 for Kendall and Distance correlation (Fig. 2). Moreover, the number of tests per infection was found strongly correlated with the CFR/IFR ratio (Fig. S4), which exceeds 1 when undetected cases are present. Larger deviation from 1 in CFR/IFR ratio indicates higher amount of undetected cases.

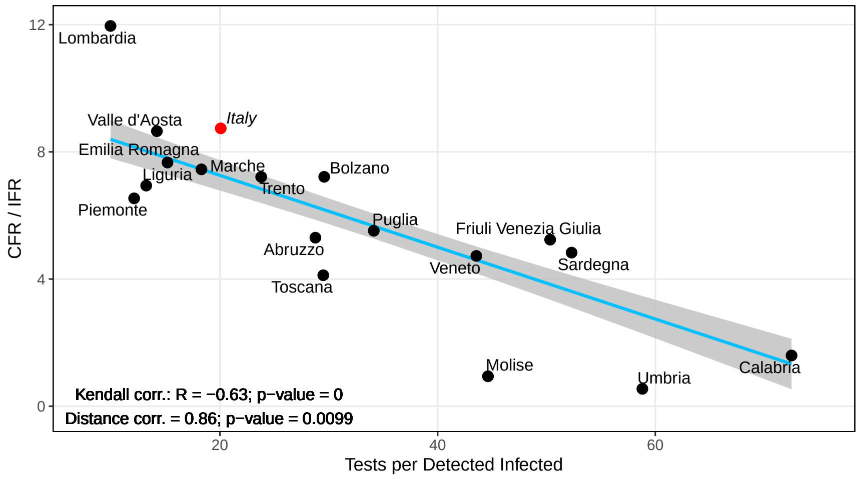


**Figure S4 Impact of the testing frequency on the regional epidemics.** Kendall and Distance correlation between the number of tests performed per infection and CFR to IFR ratio. Data are considered up to April 15$^{\text{th}}$. Light blue line: linear regression fit; gray shaded area: standard error; black dots: region-specific values; $R$: correlation coefficient; $p$: significance.

### SECIRD Models

*Reference model*

The implemented SECIRD ( Susceptible-Exposed-Carrier-Infected-Recovered-Dead) model is a deterministic ODE model with the features of SARS-CoV-2 viral infections. It distinguishes healthy individuals without immune memory of COVID-19 (susceptible, $S$), infected individuals without symptoms but not yet infectious (exposed, $E$) and infected individuals without symptoms who are infectious (carrier, $C_{\mathrm{I}}$, $C_{\mathrm{R}}$). The carriers are distinguished into asymptomatic ($C_{\mathrm{R}}$) and pre-symptomatic infected ($C_{\mathrm{I}}$), determined as $\alpha$ and $(1-\alpha)$ portion of the exposed, respectively. The pre-symptomatic infected are categorized into detected symptomatic ($I_{\mathrm{H}}$ and $I_{\mathrm{R}}$) and undetected mild-symptomatic ($I_{\mathrm{X}}$), determined as $\mu$ and $(1-\mu)$ portion of the carrier ($C_{\mathrm{I}}$). Out of the $C_{\mathrm{I}}$, $\rho$ fraction required hospitalization ($I_{\mathrm{H}}$) and $(1-\rho)$ fraction are symptomatic but recover without hospitalization ($I_{\mathrm{R}}$). Further, compartments for hospitalization ($H$) and intensive care units ($U$) were introduced to monitor the load on the healthcare system. Similarly, $\vartheta$ and $(1-\vartheta)$ represent the fraction of $H$ that requires ICU ($H_{\mathrm{U}}$) or recovered from hospital ($H_{\mathrm{R}}$), respectively. $\delta$ and $(1-\delta)$ represent the fraction of ICU who subsequently die ($U_{\mathrm{D}}$) or recover ($U_{\mathrm{R}}$). The recovered compartment ($R$) consists of recovered patients from different states of the infection. The model is summarized in Fig. 1A and details of parameter ranges used to evaluate the regional outbreak is in Table 1.

***Reference* model equations:** Equations of the *Reference* model used to fit the data (Fig. 4A and Fig. S7) and to evaluate $\mathcal{R}_t$ (Fig. 4B and Fig. S8). The description of parameters is available in Table 1. Note, $\mu = \frac{1-\bar{\mu}}{1-\alpha}$.

$$\begin{aligned}
\frac{dS}{dt} &= -R_1(t)\frac{(C_I + C_R + I_X + \beta(I_H + I_R))}{N}S \\
\frac{dE}{dt} &= R_1(t)\frac{(C_I + C_R + I_X + \beta(I_H + I_R))}{N}S - R_2 E \\
\frac{dC_I}{dt} &= (1-\alpha)\, R_2 E - R_3 C_I \\
\frac{dC_R}{dt} &= \alpha R_2 E \ - R_9 C_R \\
\frac{dI_H}{dt} &= \mu\rho(t) R_3 C_I - R_6 I_H \\
\frac{dI_R}{dt} &= \mu\,(1-\rho(t))\, R_3 C_I - R_4 I_R \\
\frac{dI_X}{dt} &= (1-\mu)\, R_3 C_I - R_4 I_X \\
\frac{dH_U}{dt} &= \vartheta(t) R_6 I_H - R_7 H_U \\
\frac{dH_R}{dt} &= (1-\vartheta(t))\, R_6 I_H - R_5 H_R \\
\frac{dU_D}{dt} &= \delta(t) R_7 H_U - R_{10} U_D \\
\frac{dU_R}{dt} &= (1-\delta(t))\, R_7 H_U - R_8 U_R \\
\frac{dR_Z}{dt} &= R_4 I_R + \ R_5 H_R + \ R_8 U_R \\
\frac{dR_X}{dt} &= R_9 C_R + \ R_4 I_X \\
\frac{dD}{dt} &= R_{10}(t) U_D
\end{aligned} \tag{1}$$

***Testing* model equations:** Equations of the *Testing* model used to quantify the impact of the undetected reduction on the hospitalized and dead compartments (Fig. 5A,B) and on the $\mathcal{R}(t)$ curve (Fig. S10). It includes an additional compartment ($I_{\mathrm{XD}}$) with respect to the basic *Reference* Model which represents the additional Detected Infected with infectivity power reduced to $\beta$. Note, $\mu = \frac{1-\bar{\mu}}{1-\alpha}$, $\mu_1(t) = \frac{\mu'(t)-\alpha}{1-\alpha}$ and $\mu_2(t) = \frac{\bar{\mu}-\mu'(t)}{1-\alpha}$ , such that $\mu_1(t) + \mu_2(t) = 1 - \mu$ .

$$
\begin{aligned}
\frac{dS}{dt} &= -R_1(t)\frac{(C_I + C_R + I_X + \beta(I_H + I_R + I_{XD}))}{N}S \\
\frac{dE}{dt} &= R_1(t)\frac{(C_I + C_R + I_X + \beta(I_H + I_R + I_{XD}))}{N}S - R_2E \\
\frac{dC_I}{dt} &= (1-\alpha)\,R_2E - R_3C_I \\
\frac{dC_R}{dt} &= \alpha R_2E \ - R_9C_R \\
\frac{dI_H}{dt} &= \mu\rho(t)R_3C_I - R_6I_H \\
\frac{dI_R}{dt} &= \mu\,(1-\rho(t))\,R_3C_I - R_4I_R \\
\frac{dI_X}{dt} &= \mu_1(t)R_3C_I - R_4I_X \\
\frac{dI_{XD}}{dt} &= \mu_2(t)R_3C_I - R_4I_{XD} \\
\frac{dH_U}{dt} &= \vartheta(t)R_6I_H - R_7H_U \\
\frac{dH_R}{dt} &= (1-\vartheta(t))\,R_6I_H - R_5H_R \\
\frac{dU_D}{dt} &= \delta(t)R_7H_U - R_{10}U_D \\
\frac{dU_R}{dt} &= (1-\delta(t))\,R_7H_U - R_8U_R \\
\frac{dR_Z}{dt} &= R_4I_{XD} + R_4I_R + \ R_5H_R + \ R_8U_R \\
\frac{dR_X}{dt} &= R_9C_R + \ R_4I_X \\
\frac{dD}{dt} &= R_{10}(t)U_D
\end{aligned}
\tag{2}
$$

***Capacity* model equations:** Equations of the *Capacity* model used to simulate the infection dynamics with limited hospital and ICU capacity (Fig. 6A and Fig. S11) and estimate its impact on the death toll (Fig. 6B). $f_{Hlim}$ and $f_{Ulim}$ functions are included in the equations to drive away the flux toward the dead compartment, from infected or hospitalized respectively, when hospital or ICU limits are reached. Note, $\mu = \frac{1-\bar{\mu}}{1-\alpha}$.

$$
\begin{aligned}
f_{Hlim} &= \left( \frac{\exp(H_U + H_R - H_{lim})^{10}}{1 + \exp(H_U + H_R - H_{lim})^{10}} \right)^{10} \Bigg|_{=0 \text{ when } H_{lim} > H_R + H_U}^{=1 \text{ when } H_{lim} < H_R + H_U} \\
f_{Ulim} &= \left( \frac{\exp(U_D + U_R - U_{lim})^{10}}{1 + \exp(U_D + U_R - U_{lim})^{10}} \right)^{10} \Bigg|_{=0 \text{ when } U_{lim} > U_D + U_R}^{=1 \text{ when } U_{lim} < U_D + U_R}
\end{aligned} \tag{3}
$$

$$
\begin{aligned}
\frac{dS}{dt} &= -R_1(t) \frac{(C_I + C_R + I_X + \beta(I_H + I_R + I_D))}{N} S \\
\frac{dE}{dt} &= R_1(t) \frac{(C_I + C_R + I_X + \beta(I_H + I_R + I_D))}{N} S - R_2 E \\
\frac{dC_I}{dt} &= (1-\alpha)\, R_2 E - R_3 C_I \\
\frac{dC_R}{dt} &= \alpha R_2 E \ - R_9 C_R \\
\frac{dI_H}{dt} &= \mu\rho(t) R_3 C_I - R_6 I_H (1 - f_{Hlim}) - R_6 I_H f_{Hlim} \\
\frac{dI_R}{dt} &= \mu\,(1-\rho(t))\, R_3 C_I - R_4 I_R \\
\frac{dI_X}{dt} &= (1-\mu)\, R_3 C_I - R_4 I_X \\
\frac{dH_U}{dt} &= \vartheta(t) R_6 I_H (1 - f_{Hlim}) - R_7 H_U \\
\frac{dH_R}{dt} &= (1-\vartheta(t)) R_6 I_H (1 - f_{Hlim}) - R_5 H_R \\
\frac{dU_D}{dt} &= \delta(t) R_7 H_U (1 - f_{Ulim}) - R_{10} U_D \\
\frac{dU_R}{dt} &= (1-\delta(t)) R_7 H_U (1 - f_{Ulim}) - R_8 U_R \\
\frac{dI_D}{dt} &= R_6 I_H f_{Hlim} - R_7 I_D \\
\frac{dR_Z}{dt} &= R_4 I_R + \ R_5 H_R + \ R_8 U_R \\
\frac{dR_X}{dt} &= R_9 C_R + \ R_4 I_X \\
\frac{dD}{dt} &= R_{10}(t) U_D + R_7 X_7 f_{Ulim} + R_7 I_D
\end{aligned} \tag{4}
$$

***TestCap* equations:** To understand the combined effect of enhanced testing and extended infrastructure upon death toll (Fig. 6), we introduced $I_{\mathrm{XD}}$ compartment into the *Capacity* Model. We sequentially drop the undetected fraction ($\bar{\mu}$) starting from its original value (Table 2) to 60% considering early (starting from one week before lockdown) and late (starting from one week post lockdown) testing strategy. Here we have used elevated hospital and ICU capacity to represent no bottleneck situation of health care system. Note, $\mu = \frac{1-\bar{\mu}}{1-\alpha}$, $\mu_1(t) = \frac{\mu'(t)-\alpha}{1-\alpha}$ and $\mu_2(t) = \frac{\bar{\mu}-\mu'(t)}{1-\alpha}$ , such that $\mu_1(t) + \mu_2(t) = 1 - \mu$ .

$$
\begin{aligned}
\frac{dS}{dt} &= -R_1(t)\frac{(C_I + C_R + I_X + \beta(I_H + I_R + I_D + I_{XD}))}{N}S \\
\frac{dE}{dt} &= R_1(t)\frac{(C_I + C_R + I_X + \beta(I_H + I_R + I_D + I_{XD}))}{N}S - R_2E \\
\frac{dC_I}{dt} &= (1-\alpha)\,R_2E - R_3C_I \\
\frac{dC_R}{dt} &= \alpha R_2E \;- R_9C_R \\
\frac{dI_H}{dt} &= \mu\rho(t)R_3C_I - R_6I_H(1 - f_{Hlim}) - R_6I_Hf_{Hlim} \\
\frac{dI_R}{dt} &= \mu\,(1-\rho(t))\,R_3C_I - R_4I_R \\
\frac{dI_X}{dt} &= \mu_1(t)R_3C_I - R_4I_X \\
\frac{dI_{XD}}{dt} &= \mu_2(t)R_3C_I - R_4I_{XD} \\
\frac{dH_U}{dt} &= \vartheta(t)R_6I_H(1 - f_{Hlim}) - R_7H_U \\
\frac{dH_R}{dt} &= (1-\vartheta(t))R_6I_H(1 - f_{Hlim}) - R_5H_R \\
\frac{dU_D}{dt} &= \delta(t)R_7H_U(1 - f_{Ulim}) - R_{10}U_D \\
\frac{dU_R}{dt} &= (1-\delta(t))R_7H_U(1 - f_{Ulim}) - R_8U_R \\
\frac{dR_Z}{dt} &= R_4I_{XD} + R_4I_R + \; R_5H_R + \; R_8U_R \\
\frac{dR_X}{dt} &= R_9C_R + \; R_4I_X \\
\frac{dD}{dt} &= R_{10}(t)U_D + R_7H_Uf_{Ulim} + R_7I_D \\
\frac{dI_D}{dt} &= R_6I_Hf_{Hlim} - R_7I_D
\end{aligned}
\tag{5}
$$

### $\mathcal{R}_t$ calculation

The basic reproduction number $R_0$ is defined as the expected number of secondary cases produced by a single infection in a completely susceptible population.

A common approach to derive this quantity for compartment models with multiple infected species is through the use of the next generation matrix method. The method was proposed by Diekmann et al. [30] and further elaborated by van den Driessche and Watmough [31]. Here we give an outline of the method for our model, but the proofs and further details can be found in [31] and [32].

First, the whole population is divided into $n$ compartments in which there are $m < n$ infected compartments which contribute to new infections. In our case, $n = 14$ and $m = 6$, with the infected compartments constituted by the exposed individuals ($E$), carriers who develop symptoms later ($C_I$), carriers who recover asymptomatically ($C_R$), undetected symptomatic people ($I_X$), identified symptomatic patients requiring hospitalization eventually ($I_H$) and identified infected individuals who recover without any hospitalization ($I_R$). Let $x_i$, $i = 1, 2, 3, \ldots, m$ be the numbers of infected individuals in the $i$-th infected compartment at time $t$. In general, we can rewrite the model equations in the form:

$$\frac{dx_i}{dt} = f_i(x) + v_i^+(x) - v_i^-(x), \tag{6}$$

where the time variation of infected people in the $i$-th compartment ($dx_i/dt$) is given by the rate of appearance of new infections in the compartment ($f_i$), plus the rate of transfer of infected individuals into the compartment ($v_i^+$), minus the rate of transfer of infected individuals out of the compartment ($v_i^-$). We now define the quantities $F$ and $V$ as the Jacobian matrices of $f_i$ and $v_i^- - v_i^+$, respectively. We evaluate these matrices at the Disease Free Equilibrium (DFE), the point at which there are no infected individuals. In our system, the $F$ and $V$ matrix have the form:

$$F = \begin{pmatrix} 0 & R_1 s_0 & R_1 s_0 & R_1 s_0 & R_1 s_0 \beta & R_1 s_0 \beta \\ 0 & 0 & 0 & 0 & 0 & 0 \\ 0 & 0 & 0 & 0 & 0 & 0 \\ 0 & 0 & 0 & 0 & 0 & 0 \\ 0 & 0 & 0 & 0 & 0 & 0 \\ 0 & 0 & 0 & 0 & 0 & 0 \end{pmatrix} \tag{7}$$

$$V = \begin{pmatrix} R_2 & 0 & 0 & 0 & 0 & 0 \\ -(1-\alpha)R_2 & R_3 & 0 & 0 & 0 & 0 \\ -\alpha R_2 & 0 & R_9 & 0 & 0 & 0 \\ 0 & -(1-\mu)R_3 & 0 & R_4 & 0 & 0 \\ 0 & -\mu\rho R_3 & 0 & 0 & R_6 & 0 \\ 0 & -\mu(1-\rho)R_3 & 0 & 0 & 0 & R_4 \end{pmatrix} \tag{8}$$

where $s_0 = S_0/N_0$, with $S_0$ and $N_0$ being the initial number of susceptible and total individuals, respectively, and the other parameters have been introduced in the main text. The DFE point is assumed to be close to the start of the epidemics and evaluating $F$ and $V$ there allows to analyze a linearization of the epidemic dynamics when the population is almost all susceptible. Once the $F$ and $V$ matrices are calculated, we define the next generation matrix $G$ as

$$G = FV^{-1}, \tag{9}$$

that is, given by the product of the matrix describing the generation of new infected individuals times the inverse of the matrix describing the net transfer of individuals across the infected compartments. We can think of the elements $g_{ij}$ of $G$ as the expected number of secondary infections of compartment $i$ caused by a single infected individual of compartment $j$. The basic reproduction number is then obtained by calculating the spectral radius of $G$, i.e. its dominant eigenvalue. For our system, this quantity reads:

$$R_0 = R_1 s_0 \left[ \frac{\alpha}{R_9} + \frac{1-\alpha}{R_3} + \frac{(1-\alpha)(1-\mu)}{R_4} + \frac{\beta(1-\alpha)(1-\rho)\mu}{R_4} + \frac{\beta(1-\alpha)\rho\mu}{R_6} \right]. \tag{10}$$

We can arrive to the same conclusion by using the formal definition of the basic reproduction number. Indeed, the chance of transmission of the disease between two individuals is directly proportional to the contact frequency ($\lambda$), i.e. how many close contacts a person makes on an average per day. If we assume that the virus has an intrinsic transmission probability of $\nu$ during each of these close contacts, the parameter $R_1$ depicts the overall chance of transmission per day due to close contacts and is given by $R_1 = \lambda\nu$. To calculate $R_0$, we also need to consider the duration for which an individual remains infectious.

At the start of a new epidemic, it is fair to assume that nobody in the population is immune to the disease and hence, the number of susceptible population $S_0$ at the beginning is same as $N_0$. Hence, the fraction of individuals who can catch infection initially is one with respect to the overall population. As the disease progresses, some people get immune to the disease following their recovery and some of them die due to the infection. This impacts the ratio of susceptible population and the overall population as both face a reduction due to recovery and death, respectively. Moreover, due to various political measures and people's responsiveness to them, the contact frequency ($\lambda$) also decreases thereby impacting $R_1$. Hence, effectively the reproduction number becomes a time varying number mainly dependent on $S(t)/N(t)$ and $R_1(t)$. If the infectivity period of an exposed person is $I_p$, it would mean

$$\mathcal{R}_{\text{t}} = R_1(t)\frac{S(t)}{N(t)}I_p, \tag{11}$$

Let's now investigate what the infectivity period is for the group of individuals of different compartments of our model. The asymptomatic carriers ($C_R$) who consist of $\alpha$ fraction of the exposed population, are capable of infecting others before they recover, i.e. for $1/R_9$ days on an average. The pre-symptomatic carriers ($C_I$), representing $(1-\alpha)$ fraction of the exposed population can infect others for an average duration of $1/R_3$ days. All the undetected symptomatic cases ($I_U$), i.e., the fraction of $(1-\alpha)(1-\mu)$ among the exposed remain infectious for a period of $1/R_4$ days following the onset of their symptoms. Similarly, the symptomatic individuals who eventually get detected but don't require hospitalization ($I_R$), i.e., the fraction of $(1-\alpha)(1-\rho)\mu$ among the exposed persons remain infectious for $1/R_4$ days. On the other hand, the detected symptomatic persons eventually requiring hospitalization ($I_H$) who represents the fraction $(1-\alpha)\rho\mu$ among the exposed people can spread the infection for $1/R_6$ days on average following their symptoms onset and before getting admitted to hospital. We assume that once somebody is admitted in a hospital cannot infect others because of suitable measures, proper isolation and protective equipment given to healthcare workers. Once somebody is detected to have an infection but still at home quarantine can still pose some risk to the susceptible population depending on how strict regulation the person follows while in home quarantine. This risk factor is represented by $\beta$.

Hence, on an average, the infectivity period of the disease is given by the summation of fractions of exposed present in a particular compartment multiplied by its average infectivity period. For the detected symptomatic cases, we also need to take into account the risk factor to spread the infection while in home quarantine. This would give an average infectivity period of

$$I_p = \frac{\alpha}{R_9} + \frac{1-\alpha}{R_3} + \frac{(1-\alpha)(1-\mu)}{R_4} + \frac{\beta(1-\alpha)(1-\rho)\mu}{R_4} + \frac{\beta(1-\alpha)\rho\mu}{R_6}, \quad (12)$$

for an exposed person in the population. This would mean that an exposed person would cause new $\mathcal{R}_\mathrm{t}$ infections in the population, where

$$\mathcal{R}(t_k) = R_1(t_k)\frac{S(t_k)}{N(t_k)}\left[\frac{\alpha}{R_9}+\frac{1-\alpha}{R_3}+(1-\mu)\frac{1-\alpha}{R_4}+\beta\mu(1-\rho(t_k))\frac{1-\alpha}{R_4}+\beta\mu\rho(t_k)\frac{1-\alpha}{R_6}\right], \quad (13)$$

where $\rho(t_k)$ denotes the hospitalized fraction of identified symptomatic cases in the $k$-th time window.

This is basically our formula for the effective reproduction rate $\mathcal{R}_\mathrm{t}$ previously derived using the next generation matrix method. Similarly, for the Testing model, we find

$$F = \begin{pmatrix} 0 & R_1 s_0 & R_1 s_0 & R_1 s_0 \beta & R_1 s_0 \beta & R_1 s_0 & R_1 s_0 \beta \\ 0 & 0 & 0 & 0 & 0 & 0 & 0 \\ 0 & 0 & 0 & 0 & 0 & 0 & 0 \\ 0 & 0 & 0 & 0 & 0 & 0 & 0 \\ 0 & 0 & 0 & 0 & 0 & 0 & 0 \\ 0 & 0 & 0 & 0 & 0 & 0 & 0 \end{pmatrix} \quad (14)$$

$$V = \begin{pmatrix} R_2 & 0 & 0 & 0 & 0 & 0 & 0 \\ -(1-\alpha)R_2 & R_3 & 0 & 0 & 0 & 0 & 0 \\ -\alpha R_2 & 0 & R_9 & 0 & 0 & 0 & 0 \\ 0 & -\mu\rho R_3 & 0 & R_6 & 0 & 0 & 0 \\ 0 & -\mu(1-\rho)R_3 & 0 & 0 & R_4 & 0 & 0 \\ 0 & -\mu_1 R_3 & 0 & 0 & 0 & R_4 & 0 \\ 0 & -\mu_2 R_3 & 0 & 0 & 0 & 0 & R_4 \end{pmatrix} \quad (15)$$

Following a similar procedure, $\mathcal{R}_{\mathrm{t}}$ of *Testing Model* can be derived as below:

$$\mathcal{R}(t_k) = R_1(t_k)\frac{S(t_k)}{N(t_k)}\Bigg[\frac{1-\alpha}{R_3} + \beta\mu\rho(t_k)\frac{1-\alpha}{R_6} + \frac{\alpha}{R_9} + \\ + \beta\mu(1-\rho(t_k))\frac{1-\alpha}{R_4} + \beta\mu_2(t_k)\frac{1-\alpha}{R_4} + \mu_1(t_k)\frac{1-\alpha}{R_4}\Bigg].$$

### Identifiability analysis

Identifiability is an important property of a mechanistic model that needs to be addressed to make a reliable prediction. Due to the practical limitations, e.g. insufficient data, measurement noise, unavailability of specific antibodies, etc., all model state variables are not observable, which limits estimation of model parameters unambiguously. Therefore, in a non-identifiable model, different sets of parameters (or states) can fit the data equally well but their prediction could be different. This has enormous implications in the context of COVID-19, especially in terms of wide variations in model predictions [33]. Structural non-identifiability [34, 35] arises due to the insufficient mapping of model states to the observables, which allows ambiguous parameters to vary without changing the observables, hence keeping $\chi^2(\theta)$ (sum of squared residuals) on a constant value. Therefore Structural non-identifiability is linked to the model structure and independent of the accuracy of available experimental data. A structurally identifiable parameter may still be practically non-identifiable and is commonly arises due to the lack of information, quality, and quantity of the considered data-sets.

To check whether our reference model is identifiable, we implemented two different strategies. We address a) structural identifiability by using synthetic outbreak data and b) practical identifiability in the setting of the current parameter estimation protocol with real data (Italy Data on Coronavirus 2020 [36]). In the synthetic outbreak method, we first randomly assign initial condition of state variables and parameter values by sampling within its range specified in Table 1. We simulate a synthetic outbreak by using this parameter set. We then use the outcome of the synthetic outbreak as model observables (dynamics of all model state variables) and asked the model to fit the data starting from randomly assigned different parameter values. This is to check whether the system reaches the same unique solution, which has been used to generate the synthetic outbreak starting from a different position in the parameter landscape. Further, structural identifiability is tested around the optimum by using the Data2Dynamics framework [37] which checks whether any directionality exists in the parameter set without sacrificing the quality of fit [38, 39]. We repeated the same procedure by using 100 different synthetic outbreak data and a typical identifiability plot is represented in Fig. S5.

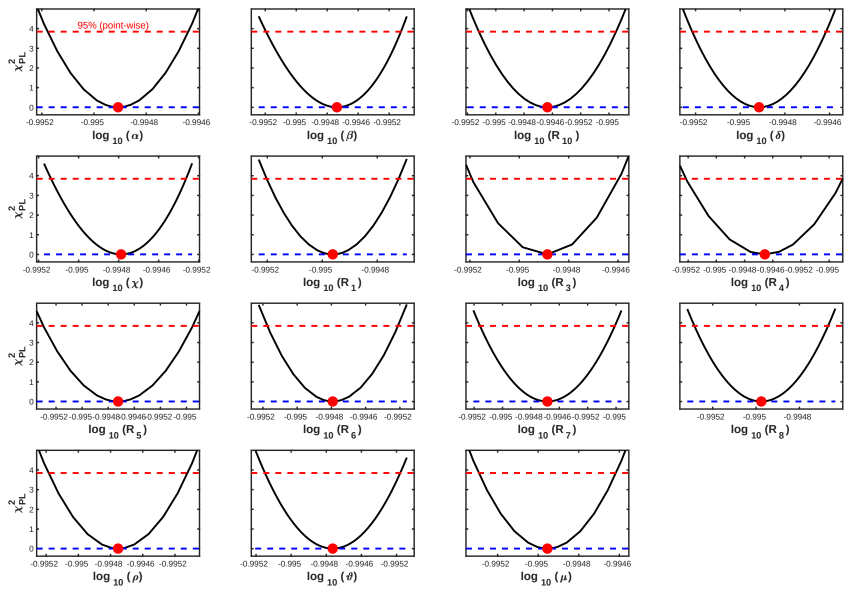


**Figure S5 Structural identifiability test.** Assessing structural identifiability of *Reference* model parameters by using synthetic outbreak data. For an identifiable parameter $\theta$, profile likelihood $\chi^2_{PL}(\theta)$ shapes like a parabola indicating a good approximation within the interval shown in each panel. Black lines indicate profile likelihood $\chi^2_{PL}(\theta)$ of parameter $\theta$ and red dot indicates the minimum value. Red lines display the threshold utilized to asses likelihood-based confidence regions [39]. A unique minimum in $\chi^2_{PL}(\theta)$ (or absence of a flat region) with respect to $\theta$ in every panel ensure that the model parameters are structurally identifiable.

To check the practical identifiability in the context of current parameter estimation setting, we first fixed all physiological parameters (see Parameterization section in Methods in the main text) and considered nationwide Italy data for the period February 24$^{th}$ to May 23$^{rd}$, 2020. We estimated behavioral parameters ($\rho, \vartheta, \delta, R_1$, $R_{10}$) by fitting the active cases, hospitalised, ICU and death data. We considered a moving time windows of 7 days and therefore checked practical identifiability of these parameters in each time window. We found that the parameters are identifiable for more than 75% of the cases. A typical result when all parameters are identifiable are represented in Fig. S6.

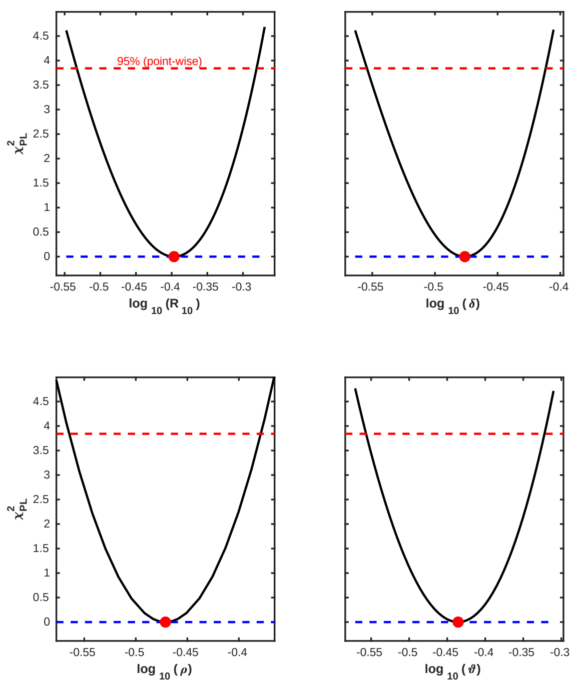


**Figure S6 Practical identifiability.** Assessing practical identifiability in the context of current parameter estimation method. Active infections, hospitalised, ICU, and death data were used as model observables to find an unique solution in $R_1$, $R_{10}$, $\rho$, $\vartheta$, and $\delta$ by keeping physiological parameters fixed. Black lines indicate profile likelihood $\chi^2_{PL}(\theta)$ of parameter $\theta$ and red dot indicates the minimum value. Red lines display the threshold utilized to asses likelihood-based confidence regions. Presence of a unique minimum in $\chi^2_{PL}(\theta)$ with respect to $\theta$ in every panel ensure that the model parameters are identifiable.

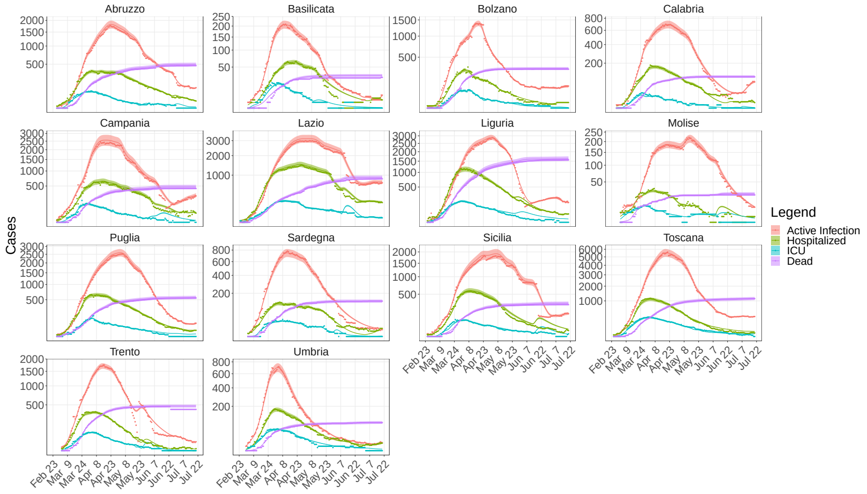


**Figure S7** **Fit up to July 23$^{\mathrm{rd}}$** Active infections, hospitalized, ICU and death data were fitted in a sliding one week time window. We present other regions that are not included in the Main text (Fig. 4A). Parameter ranges from Table 1 were used and the behavioural parameters ($R_1$, $R_{10}$, $\rho$, $\vartheta$, $\delta$) were varied in each time window (see Methods); dots: data; continuous lines: simulation results.

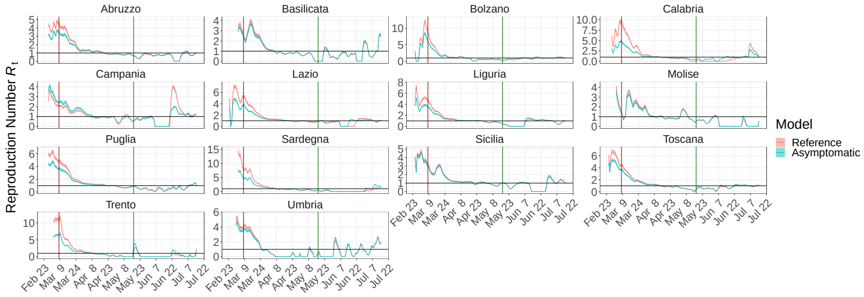


**Figure S8** $\mathcal{R}_{\mathrm{t}}$ **curves.** Boxplot of $\mathcal{R}_{\mathrm{t}}$ results from the fit of *Reference* (red) and *Asymptomatic* (blue) models. We present other regions that are not included in the Main text (Fig. 3B). Statistics performed on the $\mathcal{R}_{\mathrm{t}}$ values obtained by fitting the data with 100 perturbed parameter sets (see Methods). Vertical lines correspond to the Lockdown imposition (red) and release (green). Black dotted horizontal line represents $\mathcal{R}_{\mathrm{t}} = 1$.

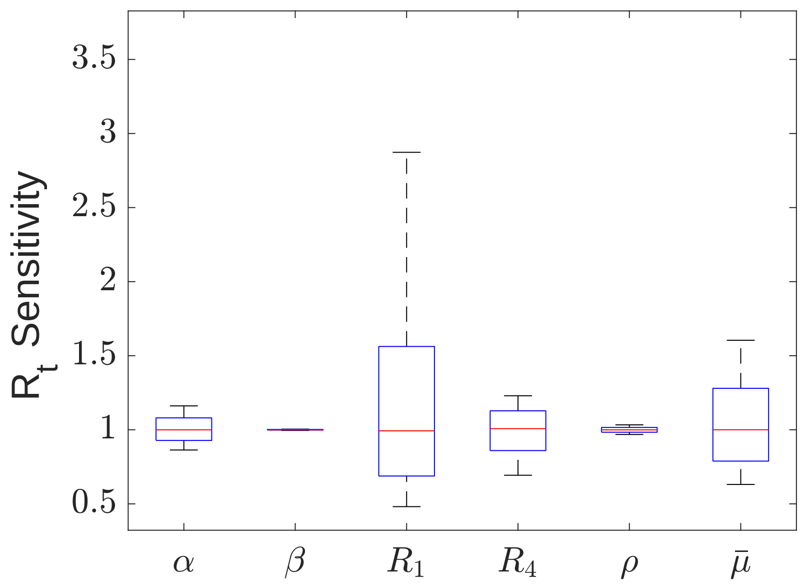


**Figure S9** $\mathcal{R}_\mathrm{t}$ **sensitivity.** Sensitivity of $\mathcal{R}_\mathrm{t}$ is performed by perturbing each parameter 20% of its reference value and by simulating the *Reference* model up to three months. Boxplot represents the variation in $\mathcal{R}_\mathrm{t}$ from its reference point. $\mathcal{R}_\mathrm{t}$ is highly sensitive to $R_1$ and $\bar{\mu}$.

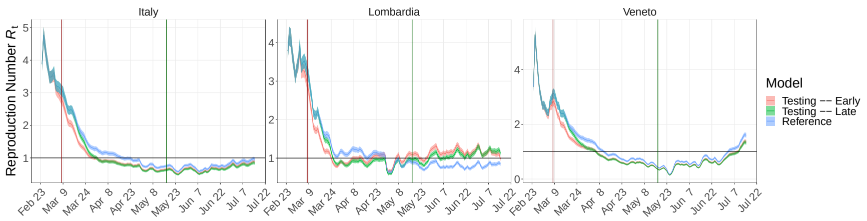


**Figure S10** **Impact of testing upon $\mathcal{R}_t$ evolution.** $\mathcal{R}_t$ curves resulting from the *Testing Model* in two evaluated scenarios: sequential decrease of undetected percentage up to 60% starting from one week before the lockdown (red) and one week post lockdown (green). $\mathcal{R}_t$ of *Reference Model* is shown in blue.

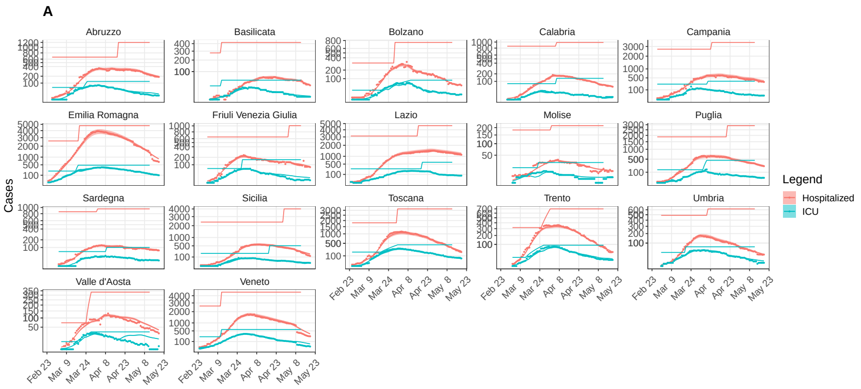


**Figure S11** ***Capacity* model fit.** Fit of active infection (not shown), hospitalized (red), ICU (iris blue) and death data (not shown) for Italian regions (except those presented in the Main text). Continuous lines: simulation results; round dots: data; Baseline capacity and linear increase in hospital (red) and ICU (iris blue) capacity (Table 3) are represented as line with respective color.

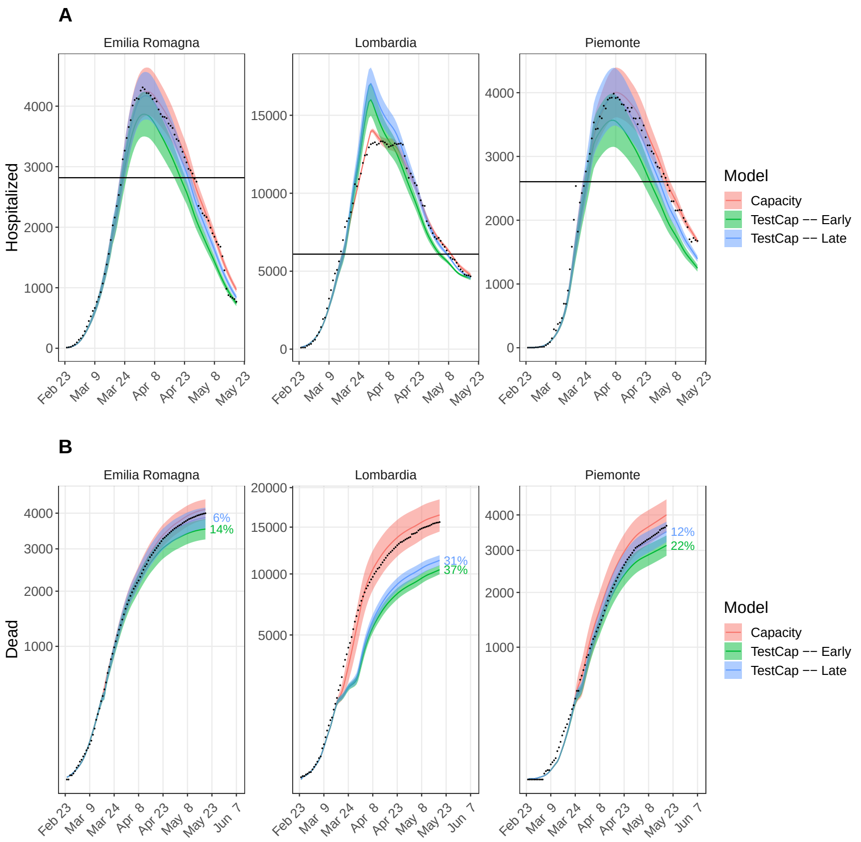


**Figure S12 Impact of 5 times more tests.** Simulation results from the *TestCap* model. Hospital and ICU capacities were set to its maximum value from the beginning and the undetected fraction ($\bar{\mu}$) is reduced starting one week before the lockdown (Early, green) or one week post lockdown (Late, light blue), resembling $\sim 5$ fold more testing than the average tests performed per day. Hospitalized is the sum of hospital and ICU patients. The reduction in dead number with respect to the data (black dots) has been reported as percentage. The black horizontal line is the capacity (hospital + ICU) before the pandemic.

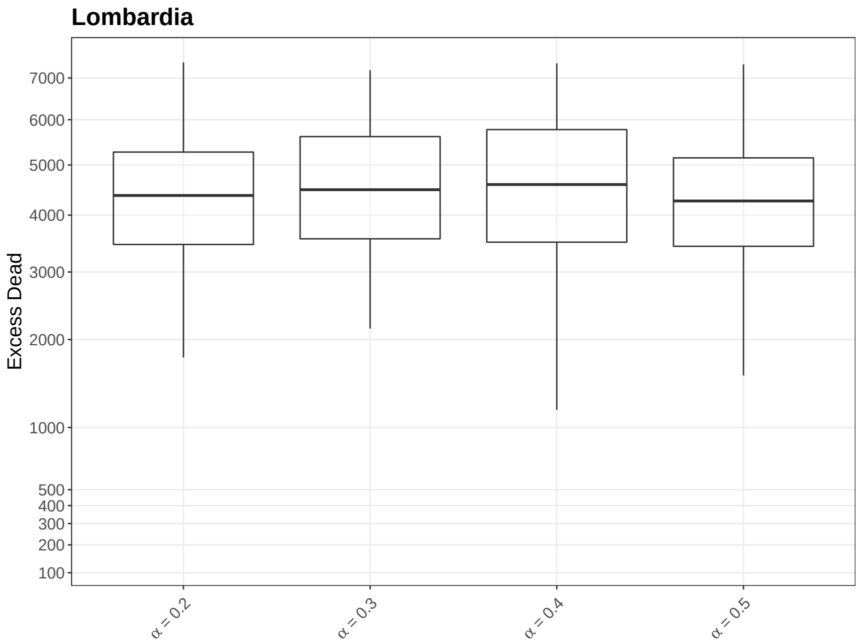


**Figure S13** **Excess Dead in Lombardia depending on $\alpha$**

**Declarations**

**Author details**

[1]Department of Systems Immunology and Braunschweig Integrated Centre of Systems Biology (BRICS), Helmholtz Centre for Infection Research, Rebenring 56, 38106, Braunschweig, Germany. [2]Institute for Biochemistry, Biotechnology and Bioinformatics, Technische Universität Braunschweig, Braunschweig, Germany. [3]Cluster of Excellence RESIST (EXC 2155), Hannover Medical School, Carl-Neuberg-Straße 1, 30625, Hannover, Germany.

**References**

1. ISTAT: Decessi e Cause di Morte: Cosa Produce l'Istat. (2020). https://www.istat.it/it/archivio/240401 (Health, Demography, Society data) Last accessed: 31 July, 2020.
2. ISTAT: ISTAT-popolazione-e-famiglie. (2019). https://www.istat.it/it/popolazione-e-famiglie?dati (Mortality data) Last accessed: 31 July, 2020.
3. Rinaldi, G., Paradisi, M.: An empirical estimate of the infection fatality rate of COVID-19 from the first Italian outbreak. medRxiv, 2020–041820070912 (2020). doi:10.1101/2020.04.18.20070912
4. Verity, R., Okell, L.C., Dorigatti, I., Winskill, P., Whittaker, C., Imai, N., Cuomo-Dannenburg, G., Thompson, H., Walker, P.G.T., Fu, H., Dighe, A., Griffin, J.T., Baguelin, M., Bhatia, S., Boonyasiri, A., Cori, A., Cucunuba, Z., FitzJohn, R., Gaythorpe, K., Green, W., Hamlet, A., Hinsley, W., Laydon, D., Nedjati-Gilani, G., Riley, S., van Elsland, S., Volz, E., Wang, H., Wang, Y., Xi, X., Donnelly, C.A., Ghani, A.C., Ferguson, N.M.: Estimates of the severity of coronavirus disease 2019: a model-based analysis. Lancet Infect Dis **20**(6), 669–677 (2020). doi:10.1016/S1473-3099(20)30243-7
5. Modi, C., Böhm, V., Ferraro, S., Stein, G., Seljak, U.: Estimating covid-19 mortality in italy early in the covid-19 pandemic. Nature communications **12**(1), 1–9 (2021)
6. Salje, H., Kiem, C.T., Lefrancq, N., Courtejoie, N., Bosetti, P., Paireau, J., Andronico, A., Hozé, N., Richet, J., Dubost, C.-L., *et al.*: Estimating the burden of sars-cov-2 in france. Science **369**(6500), 208–211 (2020)
7. Yang, W., Kandula, S., Huynh, M., Greene, S.K., Van Wye, G., Li, W., Chan, H.T., McGibbon, E., Yeung, A., Olson, D., *et al.*: Estimating the infection-fatality risk of sars-cov-2 in new york city during the spring 2020 pandemic wave: a model-based analysis. The Lancet Infectious Diseases **21**(2), 203–212 (2021)
8. Meyerowitz-Katz, G., Merone, L.: A systematic review and meta-analysis of published research data on covid-19 infection-fatality rates. International Journal of Infectious Diseases (2020)
9. Russell, T.W., Hellewell, J., Jarvis, C.I., Van Zandvoort, K., Abbott, S., Ratnayake, R., Flasche, S., Eggo, R.M., Edmunds, W.J., Kucharski, A.J., *et al.*: Estimating the infection and case fatality ratio for coronavirus disease (covid-19) using age-adjusted data from the outbreak on the diamond princess cruise ship, february 2020. Eurosurveillance **25**(12), 2000256 (2020)
10. Primi risultati dell'indagine di sieroprevalenza sul SARS-CoV-2. https://www.istat.it/it/archivio/246156 (2020)
11. Knabl, L., Mitra, T., Kimpel, J., Roessler, A., Volland, A., Walser, A., Ulmer, H., Pipperger, L., Binder, S.C., Riepler, L., Bates, K., Bandyopadhyay, A., Schips, M., Ranjan, M., Falkensammer, B., Borena, W., Meyer-Hermann, M., von Laer, D.: High SARS-CoV-2 Seroprevalence in Children and Adults in the Austrian Ski Resort Ischgl. *medRxiv*. Cold Spring Harbor Laboratory Press (2020). https://www.medrxiv.org/content/early/2020/08/22/2020.08.20.20178533.full.pdf. doi:10.1101/2020.08.20.20178533
12. Arora, R.K., Joseph, A., Van Wyk, J., Rocco, S., Atmaja, A., May, E., Yan, T., Bobrovitz, N., Chevrier, J., Cheng, M.P., et al.: SeroTracker: a global SARS-CoV-2 seroprevalence dashboard. *The Lancet. Infectious Diseases*. Elsevier (2020)
13. Team, R.C.: R: A Language and Environment for Statistical Computing. R Foundation for Statistical Computing, Vienna, Austria (2018). R Foundation for Statistical Computing
14. Youngflesh, C.: MCMCvis: Tools to visualize, manipulate, and summarize MCMC output. Journal of Open Source Software **3**(24), 640 (2018). doi:10.21105/joss.00640
15. Plummer, M., Best, N., Cowles, K., Vines, K.: CODA: Convergence Diagnosis and Output Analysis for MCMC. R News **6**(1), 7–11 (2006)
16. Sturtz, S., Ligges, U., Gelman, A.: R2WinBUGS: A Package for Running WinBUGS from R. Journal of Statistical Software **12**(3), 1–16 (2005)
17. Baud, D., Qi, X., Nielsen-Saines, K., Musso, D., Pomar, L., Favre, G.: Real estimates of mortality following COVID-19 infection. Lancet Infect Dis **20**(7), 773 (2020). doi:10.1016/S1473-3099(20)30195-X
18. Kim, D.H., Choe, Y.J., Jeong, J.Y.: Understanding and Interpretation of Case Fatality Rate of Coronavirus Disease 2019. J Korean Med Sci **35**(12), 137 (2020). doi:10.3346/jkms.2020.35.e137
19. Nishiura, H., Linton, N.M., Akhmetzhanov, A.R.: Serial interval of novel coronavirus (COVID-19) infections. Int J Infect Dis **93**, 284–286 (2020). doi:10.1016/j.ijid.2020.02.060
20. Zhou, X., Li, Y., Li, T., Zhang, W.: Follow-up of asymptomatic patients with SARS-CoV-2 infection. Clin Microbiol Infect **26** (2020). doi:10.1016/j.cmi.2020.03.024
21. Chen, N., Zhou, M., Dong, X., Qu, J., Gong, F., Han, Y., Qiu, Y., Wang, J., Liu, Y., Wei, Y., Xia, J., Yu, T., Zhang, X., Zhang, L.: Epidemiological and clinical characteristics of 99 cases of 2019 novel coronavirus pneumonia in Wuhan, China: a descriptive study. Lancet **395**(10223), 507–513 (2020). doi:10.1016/S0140-6736(20)30211-7
22. Chan, J.F., Yuan, S., Kok, K.H., To, K.K., Chu, H., Yang, J., Xing, F., Liu, J., Yip, C.C., Poon, R.W., Tsoi, H.W., Lo, S.K., Chan, K.H., Poon, V.K., Chan, W.M., Ip, J.D., Cai, J.P., Cheng, V.C., Chen, H., Hui, C.K., Yuen, K.Y.: A familial cluster of pneumonia associated with the 2019 novel coronavirus indicating person-to-person transmission: a study of a family cluster. Lancet **395**(10223), 514–523 (2020). doi:10.1016/S0140-6736(20)30154-9
23. Guan, W.J., Ni, Z.Y., Hu, Y., Liang, W.H., Ou, C.Q., He, J.X., Liu, L., Shan, H., Lei, C.L., Hui, D.S.C., Du, B., Li, L.J., Zeng, G., Yuen, K.Y., Chen, R.C., Tang, C.L., Wang, T., Chen, P.Y., Xiang, J., Li, S.Y., Wang, J.L., Liang, Z.J., Peng, Y.X., Wei, L., Liu, Y., Hu, Y.H., Peng, P., Wang, J.M., Liu, J.Y., Chen, Z., Li, G.,

Zheng, Z.J., Qiu, S.Q., Luo, J., Ye, C.J., Zhu, S.Y., Zhong, N.S., China Medical Treatment Expert Group for, C.: Clinical Characteristics of Coronavirus Disease 2019 in China. N Engl J Med **382**(18), 1708–1720 (2020). doi:10.1056/NEJMoa2002032
24. Huang, C., Wang, Y., Li, X., Ren, L., Zhao, J., Hu, Y., Zhang, L., Fan, G., Xu, J., Gu, X., Cheng, Z., Yu, T., Xia, J., Wei, Y., Wu, W., Xie, X., Yin, W., Li, H., Liu, M., Xiao, Y., Gao, H., Guo, L., Xie, J., Wang, G., Jiang, R., Gao, Z., Jin, Q., Wang, J., Cao, B.: Clinical features of patients infected with 2019 novel coronavirus in Wuhan, China. Lancet **395**(10223), 497–506 (2020). doi:10.1016/S0140-6736(20)30183-5
25. Li, Q., Guan, X., Wu, P., Wang, X., Zhou, L., Tong, Y., Ren, R., Leung, K.S.M., Lau, E.H.Y., Wong, J.Y., Xing, X., Xiang, N., Wu, Y., Li, C., Chen, Q., Li, D., Liu, T., Zhao, J., Liu, M., Tu, W., Chen, C., Jin, L., Yang, R., Wang, Q., Zhou, S., Wang, R., Liu, H., Luo, Y., Liu, Y., Shao, G., Li, H., Tao, Z., Yang, Y., Deng, Z., Liu, B., Ma, Z., Zhang, Y., Shi, G., Lam, T.T.Y., Wu, J.T., Gao, G.F., Cowling, B.J., Yang, B., Leung, G.M., Feng, Z.: Early Transmission Dynamics in Wuhan, China, of Novel Coronavirus-Infected Pneumonia. N Engl J Med **382**(13), 1199–1207 (2020). doi:10.1056/NEJMoa2001316
26. Villa, M., Myers, J.F., Turkheimer, F.: COVID-19: Recovering estimates of the infected fatality rate during an ongoing pandemic through partial data. medRxiv (2020). doi:10.1101/2020.04.10.20060764. https://www.medrxiv.org/content/early/2020/04/14/2020.04.10.20060764.full.pdf
27. ISTAT: IMPATTO DELL'EPIDEMIA COVID-19 SULLA MORTALITÀ TOTALE DELLA POPOLAZIONE RESIDENTEPRIMO TRIMESTRE. (2020). `https://www.istat.it/it/files/2020/05/Rapporto_Istat_ISS.pdf` Last accessed: 31 July, 2020
28. Grasselli, G., Pesenti, A., Cecconi, M.: Critical Care Utilization for the COVID-19 Outbreak in Lombardy, Italy: Early Experience and Forecast During an Emergency Response. JAMA **323**(16), 1545–1546 (2020). doi:10.1001/jama.2020.4031
29. Vollmer, M., Mishra, S., Juliette, H., et al.: Report 20: Using mobility to estimate the transmission intensity of COVID-19 in Italy: a subnational analysis with future scenarios. *Imperial College London*. Imperial College London (2020)
30. Diekmann, O., Heesterbeek, J.A., Metz, J.A.: On the definition and the computation of the basic reproduction ratio R0 in models for infectious diseases in heterogeneous populations. J Math Biol **28**(4), 365–382 (1990). doi:10.1007/BF00178324. Accessed 2020-04-07
31. van den Driessche, P., Watmough, J.: Reproduction numbers and sub-threshold endemic equilibria for compartmental models of disease transmission. Math Biosci **180**, 29–48 (2002). doi:10.1016/S0025-5564(02)00108-6. Accessed 2020-06-17
32. Van den Driessche, P., Watmough, J.: Further notes on the basic reproduction number. In: Mathematical Epidemiology, pp. 159–178. Springer, ??? (2008)
33. Roda, W.C., Varughese, M.B., Han, D., Li, M.Y.: Why is it difficult to accurately predict the covid-19 epidemic? Infectious Disease Modelling **5**, 271–281 (2020)
34. Walter, E.: Identifiability of Parametric Models. Elsevier, ??? (2014)
35. Cobelli, C., Distefano 3rd, J.J.: Parameter and structural identifiability concepts and ambiguities: a critical review and analysis. American Journal of Physiology-Regulatory, Integrative and Comparative Physiology **239**(1), 7–24 (1980)
36. COVID-19 Italia - Monitoraggio situazione. `https://github.com/pcm-dpc/COVID-19` (2020)
37. Raue, A., Steiert, B., Schelker, M., Kreutz, C., Maiwald, T., Hass, H., Vanlier, J., Tonsing, C., Adlung, L., Engesser, R., Mader, W., Heinemann, T., Hasenauer, J., Schilling, M., Hofer, T., Klipp, E., Theis, F., Klingmuller, U., Schoberl, B., Timmer, J.: Data2Dynamics: a modeling environment tailored to parameter estimation in dynamical systems. Bioinformatics **31**(21), 3558–60 (2015). doi:10.1093/bioinformatics/btv405
38. Raue, A., Kreutz, C., Maiwald, T., Bachmann, J., Schilling, M., Klingmüller, U., Timmer, J.: Structural and practical identifiability analysis of partially observed dynamical models by exploiting the profile likelihood. Bioinformatics **25**(15), 1923–1929 (2009). doi:10.1093/bioinformatics/btp358. https://academic.oup.com/bioinformatics/article-pdf/25/15/1923/16889623/btp358.pdf
39. Raue, A., Kreutz, C., Maiwald, T., Klingmüller, U., Timmer, J.: Addressing parameter identifiability by model-based experimentation. IET systems biology **5**(2), 120–130 (2011)
